# Predicting Nonsense-mediated mRNA Decay from Splicing Events in Sepsis using RNA-Sequencing Data

**DOI:** 10.1101/2025.03.31.25324958

**Authors:** Jaewook Shin, Alger M. Fredericks, Brandon E. Armstead, Alfred Ayala, Maya Cohen, William G. Fairbrother, Mitchell M. Levy, Kwesi K. Lillard, Emanuele Raggi, Gerard J. Nau, Sean F. Monaghan

**Author notes:** **Corresponding author:** Sean F. Monaghan, MD. Address: 593 Eddy Street, Middle House 211, Providence, RI 02903.

## Abstract

Alternative splicing (AS) and nonsense-mediated mRNA decay (NMD) are highly conserved cellular mechanisms that modulate gene expression. Here we introduce NMD pipeline that computes how splicing events introduce premature termination codons to mRNA transcripts via frameshift, then predicts the rate of PTC-dependent NMD. We utilize whole blood, deep RNA-sequencing data from critically ill patients to study gene expression in sepsis. Statistical significance was determined as adjusted p value < 0.05 and |log2foldchange| > 2 for differential gene expression and probability >= 0.9 and |DeltaPsi| > 0.1 for AS. NMD pipeline was developed based on AS data from Whippet. We demonstrate that the rate of NMD is higher in sepsis and deceased groups compared to control and survived groups, which signify purposeful downregulation of transcripts by AS-NMD or aberrant splicing due to altered physiology. Predominance of non-exon skipping events was associated with disease and mortality states. The NMD pipeline also revealed proteins with potential novel roles in sepsis. Together, these results emphasize the utility of NMD pipeline in studying AS-NMD along with differential gene expression and discovering potential protein targets in sepsis.

## Introduction

Alternative splicing (AS) and nonsense-mediated mRNA decay (NMD) are crucial molecular processes that modulate gene expression (1, 2). AS contributes to protein diversity in higher eukaryotes and close to 95% of multiexon genes are estimated to undergo AS in major human tissues (3). NMD is a highly conserved surveillance mechanism that eliminates mRNAs based on multiple criteria, most commonly when premature termination codons (PTC) are located more than 50-55 nucleotide upstream of the final exon junction (4). Since splicing events can generate PTCs via frameshift, AS-NMD may downregulate genes or decay aberrant transcripts (5) thereby fine-tuning gene expression and maintaining cellular homeostasis (6). Recently, AS and NMD have been indicated in the pathogenesis of malignancy (7–9), critical illness (10–12), and in the changes seen in gene expression due to altered physiological states such as hypoxia, acidosis, and temperature (13, 14).

Sepsis is a leading cause of mortality worldwide responsible for up to 1 in 5 deaths (15), and occurs when infection causes a dysregulated host response leading to life-threatening organ dysfunction (16). While earlier diagnosis and guidelines have shown some benefit, current understanding of sepsis pathogenesis has not substantially improved patient outcomes (17). Despite the efforts to understand sepsis-induced cellular and subcellular dysfunction by characterizing gene expression profiles (18–21), none have translated to bedside treatments (22). Given that differential gene expression (DGE) alone has been ineffective, studying AS-NMD in conjunction can delineate how intermediate steps of gene expression affect downstream proteins.

Here, we demonstrate a computational pipeline that predicts NMD from AS data generated from patients’ whole blood, deep RNA sequencing (RNA-Seq) data. This pipeline computes how splicing events generate PTCs via frameshift and affect protein levels and how such prediction identifies proteins with potential novel roles in sepsis and mortality. For this work, we focus on NMD that is PTC-dependent and targets PTCs located 50-55 bp upstream of final exon junction. We hypothesize that there will be more splicing events leading to NMD in sepsis and deceased group due to altered physiology that either purposefully or aberrantly decays select transcripts.

## Results

### Patient characteristics of control versus sepsis groups and survived versus deceased groups

A total of 43 critically ill patients with sepsis and 6 without sepsis were studied. Among sepsis patients, mortality rate was 34.9% with 25 patients who survived and 18 who deceased. The sepsis group was older than the controls (43.8±18 years vs. 63.2±14.9 years, p=0.049) but did not have significantly different percentages of male (33.3% vs. 55.8%, p=0.55) or non-Caucasians (50% vs. 27.9%, p=0.53). Similarly, patients’ age, sex, and race did not differ based on mortality. Compared to control, sepsis group had statistically significantly higher rate of shock (0% vs. 53.5%, p=0.02), longer median ICU length of stay (LOS) (1.3 vs. 2.2 days; p<0.01), and longer median hospital LOS (2.8 vs. 9.2 days, p<0.01). Survived and deceased groups had similar rate of shock (56% vs. 50%, p=0.76), SOFA score (4.5 vs. 6, p=0.29), ICU (2 vs. 4.4 days, p=0.05) and hospital LOS (9.5 vs. 7.8 days, p=0.61) (Table 1).

**Table 1.** Patient demographics, clinical outcomes, and RNA-Seq data in control vs sepsis and survived vs deceased. Mortality refers to the rate of in-hospital death. SOFA score refers to Sequential Organ Failure Assessment (SOFA) score. Total reads refer to total number of RNA-Seq reads yielded from each patient group, with respective percentages of reads mapped to human genome (“mapped”) and not mapped to human genome (“unmapped”).

|  | Control | Sepsis | p value | Survived | Deceased | p value |
| --- | --- | --- | --- | --- | --- | --- |
| **Sample size (N)** | 6 | 43 | -- | 25 | 18 | -- |
| **Age, mean (years)** | 43.8 ± 18.4 | 63.2 ± 14.9 | 0.049 | 63.9 ± 16.3 | 61.9 ± 12.2 | 0.66 |
| **Male (%)** | 2 (33.3%) | 24 (55.8%) | 0.55 | 13 (46.4%) | 11 (73.3%) | 0.17 |
| **Non-Caucasian (%)** | 3 (50%) | 12 (27.9%) | 0.53 | 8 (28.6%) | 4 (26.7%) | 0.84 |
| **Mortality (%)** | 0 (0%) | 15 (34.9%) | 0.16 | -- | -- | -- |
| **Shock (%)** | 0 (0%) | 23 (53.5%) | 0.02 | 14 (56%) | 9 (50%) | 0.76 |
| **SOFA (score)** | 4 | 5.5 | 0.06 | 4.5 | 6 | 0.29 |
| **ICU stay, median (days)** | 1.3 | 2.2 | <0.01 | 2 | 4.4 | 0.05 |
| **Hospital stay, median (days)** | 2.8 | 9.2 | <0.01 | 9.5 | 7.8 | 0.61 |
| **Total reads, mean (N)** | 120,199,186 | 116,110,683 | 0.57 | 118,008,653 | 112,567,805 | 0.14 |
| **Mapped, mean, N (%)** | 96,239,315 (80.1%) | 92,755,097 (79.9%) | 0.47 | 92,177,756 (78.2%) | 93,832,802 (83.4%) | 0.63 |
| **Unmapped, median, N (%)** | 21,895,386 (18.2%) | 19,715,777 (17%) | 0.99 | 24,944,197 (21.8%) | 17,869,942 (16.6%) | <0.01 |

### Whole blood, non-poly(A) selected, deep RNA-Seq pipeline enables DGE and AS study

Next, deep RNA-Seq of patients’ whole blood yielded a total of 11.1 billion reads across all samples. Non-poly(A) tail selection facilitated the comprehensive AS analysis since all transcripts with and without poly(A) tail were included. Of the total reads, 8.8 billion reads were mapped to human genome with 2.3 billion reads unmapped (Figure 1A). Mean total RNA-Seq reads per sample were similar between control and sepsis groups (120 million vs. 116 million, p=0.57) and survived and deceased groups (118 million vs. 112 million, p=0.14), confirming the efficacy of deep RNA sequencing yielding at least 100 million reads per sample (Table 1). The mapped reads were then processed for downstream analyses including DGE analysis of 17,043 genes and AS studies with 220,779 splicing events (Figure 1A).

**Fig. 1.**
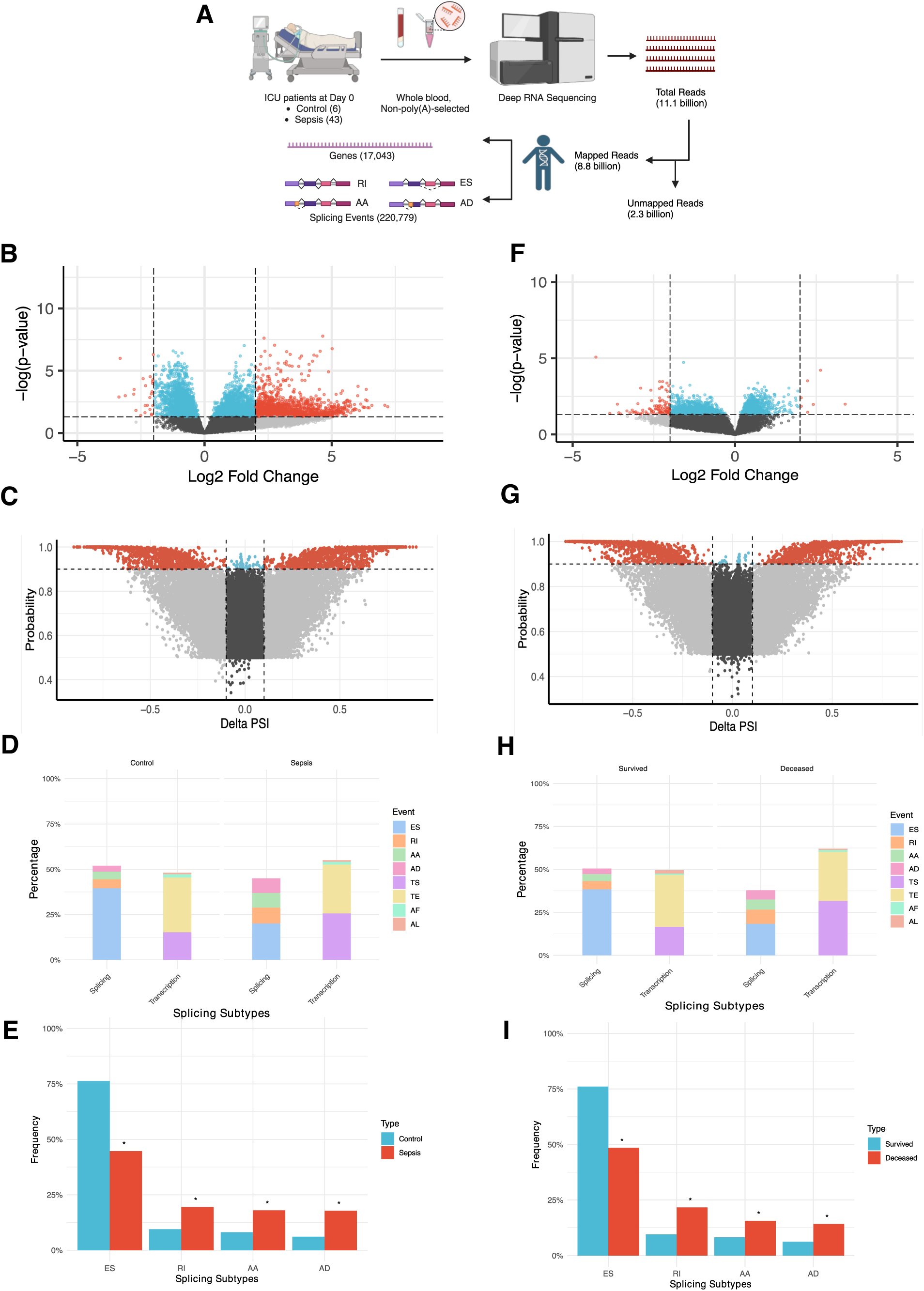
Differential gene expression (DGE) and alternative splicing (AS) data in control versus sepsis (Fig. 1B-E) and survived versus deceased groups (Fig. 1F-I). **(A)** Diagram describing RNA-Seq workflow from ICU patients to their DGE and AS information. Created in https://BioRender.com. **(B)** Volcano plot showing DGE analysis of significantly up- or down-regulated genes in control vs sepsis based on adjusted p value and Log2 Fold Change (Log2 FC) (red), adjusted p value alone (blue), Log2 FC alone (grey), and not statistically significant (black). **(C)** Volcano plot showing differential splicing analysis of significantly more or less frequent splicing events in control vs. sepsis based on probability and Delta Percent-Spliced In (Delta PSI) (red), probability alone (blue), Delta PSI alone (grey), and not statistically significant (black). **(D)** Proportion of each subtype out of all splicing events in control vs. sepsis groups then categorized into “Splicing” and “Transcription-related” groups. **(E)** Frequency of each of the four splicing events (from “Splicing” group in Fig. 3D) in percentage in control vs sepsis. **(F)** Volcano plot showing DGE analysis of significantly up- or down-regulated genes in survived vs deceased with same color and statistical depictions as Fig. 1B. **(G)** Volcano plot showing differential splicing analysis of significantly more or less frequent splicing events in survived vs. deceased with same color and statistical depiction as Fig. 1C. **(H)** Proportion of each subtype out of all splicing events in survived vs. deceased groups then categorized into “Splicing” and “Transcription-related groups. **(I)** Frequency of each of the four splicing events (from “Splicing” group in Fig. 3H) in percentage in survived vs deceased.

### More upregulated genes but similar rate of AS in sepsis compared to control group

First, we examined the DGE and AS profiles of control and sepsis groups. Of the 17,043 genes differentially expressed between the two groups, 1,349 genes (7.9%) were significantly differentially expressed with 1,325 upregulated (98.2%) and 24 (1.8%) downregulated in sepsis showing that more genes analyzed were highly expressed in sepsis (Figure 1B, S1). There were 220,779 events differentially spliced in control versus sepsis, with 2,158 splicing events (1%) significantly differentially frequent. Of these, 1,014 events (47%) were more frequent in sepsis and 1,144 events (53%) less frequent, highlighting that splicing events were more evenly split in contrast to DGE results (Figure 1C, S2). We then categorized 220,779 splicing events into subtypes so that splicing events mediated exclusively by the splicing machinery could be distinguished from alternative transcription sites. These subtypes were then compared between all splicing events (effectively representing the “Control” group) and significant differential splicing events in sepsis (“Sepsis” group). The results showed that alternative transcription events represented 48.1% and 55% in each group, with transcription start (TS) and end (TE) constituting the highest percentages (94.5%, 95.6%), whereas splicing events were 51.9% and 45% in each group with exon skipping events (ES) constituting the highest percentages (76.3%, 44.7%) (Figure 1D, Table S1). Then, we selected four splicing events – exon skipping (ES), retained intron (RI), alternative acceptor (AA), and alternative donor (AD) – and compared their frequency between control and sepsis groups, showing that ES was significantly less frequent in sepsis (76.3% vs. 44.7%, p<0.001) while other events were significantly more frequent in sepsis (RI 9.5% vs. 19.5%, p<0.001; AA 8.1% vs. 18%, p<0.001; AD 6.1% vs. 17.8%, p<0.001), demonstrating that predilection for non-ES splicing events were present in sepsis (Figure 1E, Table S2).

### More downregulated genes but similar rate of AS in deceased compared to survived group

Next, we examined the DGE and AS profiles of sepsis patients based on their mortality status. Of the 16,837 genes differentially expressed between survived and deceased groups, 118 genes (0.7%) were significantly differentially expressed with 7 upregulated (5.9%) and 111 (94.1%) downregulated in sepsis showing that more genes analyzed were significantly less expressed in deceased group (Figure 1F, S3). There were 233,753 differential splicing events with 2,282 significantly differential splicing events (1%). Of these, 1,172 events (47%) were more frequent in deceased group and 1,110 events (53%) less frequent, again highlighting the similar degree of splicing in contrast to DGE results (Figure 1G, S4). We then categorized 233,753 splicing events into splicing and alternative transcription subtypes. The results were consistent with control versus sepsis analysis, showing that alternative transcription events represented 49.5% and 62.1% in each group, with TS and TE constituting the highest percentages (94.6%, 97%), whereas splicing events were 50.5% and 37.9% in each group with ES being the highest percentages (76.1%, 48.5%) (Figure 1H, Table S3). The frequency of all differential splicing events (“Survived” group) and significant differential splicing events (“Deceased” group) showed that ES was significantly less frequent in deceased (76.1% vs. 48.5%, p<0.001) while all other events were significantly more frequent in deceased (RI 9.5% vs. 21.7%, p<0.001; AA 8.2% vs. 15.6%, p<0.001; AD 6.2% vs. 14.2%, p<0.001), demonstrating a similar pattern between sepsis and deceased groups regarding splicing events (Figure 1I, Table S4).

### Developing a computational pipeline to predict NMD with splicing data from Whippet

To study NMD, we reasoned that AS data from Whippet (23) can show if and how many PTCs would be generated from each splicing event. The scope of the pipeline was to predict PTC generation by 4 splicing events (ES, RI, AA, AD) based on the established principle that the presence of PTC is expected to elicit NMD (13). Thus, we selected key splicing information, such as Ensembl ID, splicing event type, splicing coordinate, node, and strand to model nucleotide sequences of mature transcripts resulting from each splicing event and to identify how frameshifts would generate PTCs upstream of 50-55 base pairs from the final exon junction in accordance with one of the accepted prerequisites for NMD (4). As a result, our NMD pipeline yields the following outputs: the predicted frame of each transcript, the number and location of all PTCs generated per frame, and predicted NMD true or false based on the predicted frame and PTC location (Figure 2A, Supplementary Text).

**Fig. 2.**
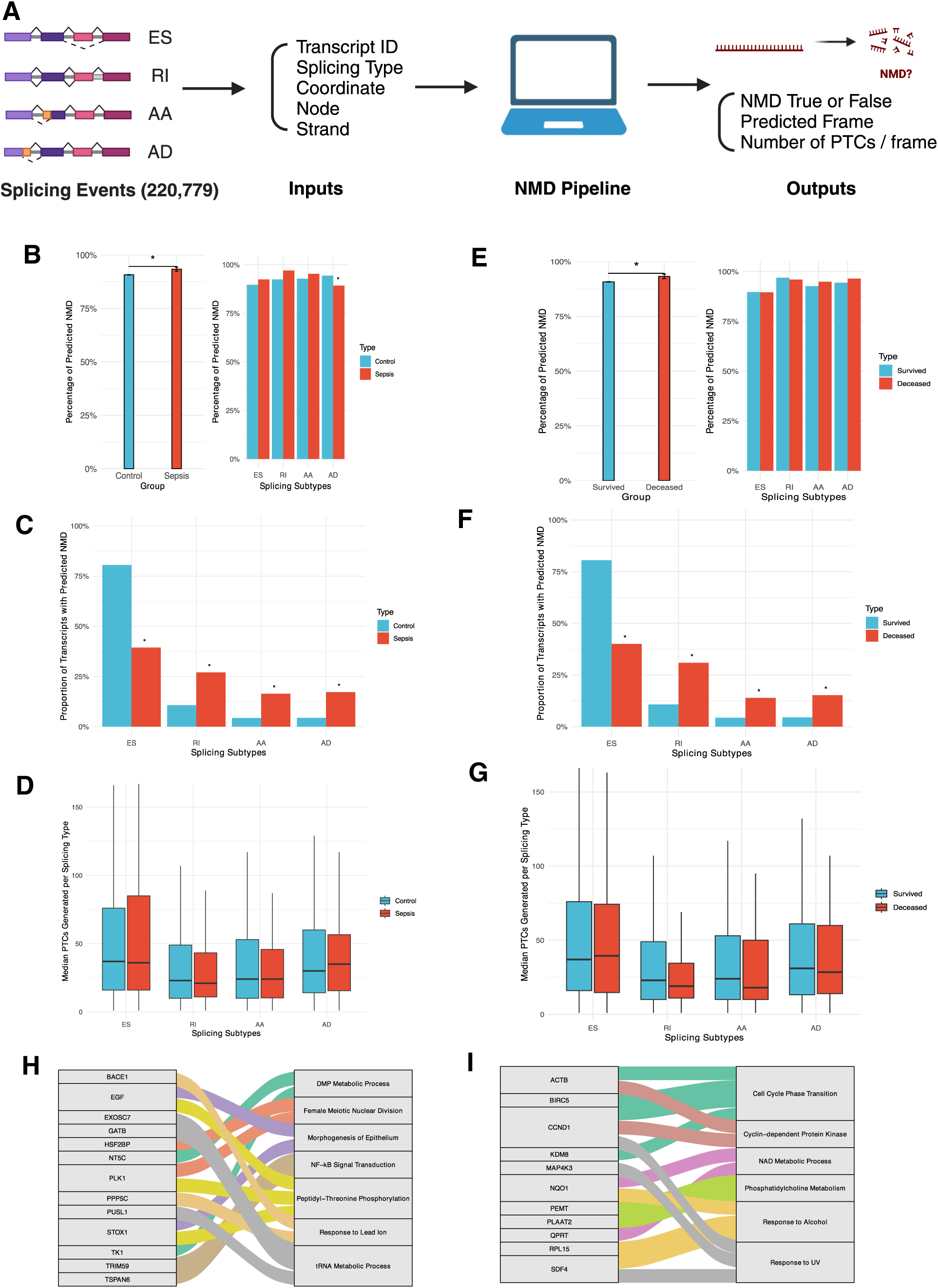
Alternative splicing (AS) and nonsense-mediated mRNA decay (NMD) data in control versus sepsis (Fig. 1B-D, H) and survived versus deceased groups (Fig. 1E-G, I). **(A)** Diagram describing the development of NMD pipeline from Whippet AS data to NMD outputs. Created in https://BioRender.com. **(B)** Bar graph showing the percentage of splicing events predicted to induce NMD in control vs sepsis (left) and the percentage of predicted NMD stratified by splicing subtypes (right). **(C)** Proportion of splicing events of transcripts predicted to cause NMD per each splicing subtype in control vs sepsis. **(D)** Box plot showing the median number of premature termination codons (PTCs) generated per splicing subtype in control vs sepsis. **(E)** Bar graph showing the percentage of splicing events predicted to induce NMD in survived vs deceased (left) and the percentage of predicted NMD stratified by splicing subtypes (right). **(F)** Proportion of splicing events of transcripts predicted to cause NMD per each splicing subtype in survived vs deceased. **(G)** Box plot showing the median number of premature termination codons (PTCs) generated per splicing subtype in survived vs deceased. **(H)** Sankey diagram showing all the genes with p < 0.01 in GO Enrichment Analysis and their respective biological processes in control vs sepsis. **(I)** Sankey diagram showing all the genes with p < 0.01 in GO Enrichment Analysis and their respective biological processes in survived vs deceased.

### Higher rate of predicted NMD in sepsis group from non-ES splicing events

Utilizing the NMD pipeline, we compared the overall rate of NMD in sepsis compared to control. We processed all splicing events and those significant in sepsis through the pipeline and found a significantly higher rate of NMD predicted to occur from splicing events in sepsis compared to control (90.7% vs. 93.3%, p=0.03) and each splicing subtype predicted to induce NMD at a similar rate, except for AD (94.4% vs. 89.2%, p=0.03) (Figure 2B, Table S5). ES was the most common splicing event predicted to cause NMD though less frequently in sepsis (80.5%vs.39.4%, p<0.001). There were significantly higher percentage of RI, AA, and AD splicing events predicted to induce NMD in sepsis (RI 10.7% vs. 27%, p<0.001; AA 4.3% vs. 16.4%, p<0.001; AD 4.4% vs. 17.2%, p<0.001) (Figure 2C, Table S6). While ES generated the highest median number of PTCs, none of the splicing events in sepsis introduced significantly different amount of PTCs compared to all splicing events (ES 37 vs. 36, p=0.51; RI 23 vs. 21, p=0.68; AA 24 vs. 24, p=0.74; AD 30 vs. 35, p=0.43) showing that the number of PTCs generated is not correlated with a higher rate of NMD seen in sepsis (Figure 2D, Table S7).

### Higher rate of predicted NMD in deceased group from non-ES splicing events

We also studied whether the rate of NMD would be significantly different based on mortality and found a significantly higher rate of NMD predicted to occur in deceased group (90.8% vs. 93.3%, p=0.04) and each splicing subtype predicted to induce NMD at a similar rate (Figure 2E, Table S8). Similarly, ES most predicted NMD though less frequently in deceased (80.5% vs. 40%, p<0.001). Again, there were significantly higher percentages of RI, AA, and AD events predicted to induce NMD in deceased (RI 10.7% vs. 30.9%, p<0.001; AA 4.3% vs. 13.9%, p<0.001; AD 4.5% vs. 15.2%, p<0.001) (Figure 2F, Table S9). ES accounted for the highest median number of PTCs generated but splicing events did not introduce meaningfully different median number of PTCs, consistent with control vs sepsis analysis that the number of PTCs generated doees not correlate with the higher rate of NMD in deceased group (ES 37 vs. 39.5, p=0.73; RI 23 vs. 19, p=0.07; AA 24 vs. 18, p=0.47; AD 31 vs. 28.5, p=0.58) (Figure 2G, Table S10).

### NMD pipeline can identify proteins with potential novel roles in sepsis and mortality

We then examined whether splicing events not predicted to cause NMD, thus preserving certain transcripts, could identify proteins with potentially novel roles in sepsis and mortality. We performed GO enrichment analysis of splicing events yielding predicted NMD result as “false” (NMD-F) then filtered by p<0.01. In control vs. sepsis, 45 splicing events were NMD-F and 13 genes were identified as highly likely to be relevant to 7 biological processes, including essential nucleic acid metabolism (e.g. DMP, tRNA), cell division and development (e.g. meiosis, epithelium), inflammation (e.g. NF-kB, phosphorylation), and response to stressor (e.g. lead) (Figure 2H, Table S11). In survived vs. deceased, 39 splicing events were NMD-F and 11 genes were identified as highly likely to be relevant to 6 biological processes, including essential nucleic acid metabolism (e.g. NAD), cell division and development (e.g. cell cycle, CDK), inflammation (e.g. phosphatidylcholine), and response to stressor (e.g. alcohol, UV) (Figure 2I, Table S12). Thus, NMD-F from the NMD pipeline can identify proteins with known sepsis-relevant or essential cellular functions (i.e. inflammation, nucleic acid and cell metabolism) and potential novel roles (i.e. response to alcohol, UV) not previously known in sepsis.

### Plasma grancalcin concentration is higher in sepsis as predicted by NMD pipeline

Finally, we used a representative protein to test NMD pipeline predictions and evaluate the impact of NMD on protein abundance. Grancalcin (GCA) was chosen since it was the only gene with significant differential splicing events in both sepsis and deceased groups, each with 2,656 and 866 events, and with one of the highest RNA-Seq read counts overall in both groups (Figure 3A, Supplementary Text). Between control and sepsis, GCA was not significantly differentially expressed based on DGE, less frequently spliced in sepsis based on Whippet, and NMD predicted true based on the NMD pipeline. Thus, higher protein level was predicted in sepsis due to fewer splicing events inducing NMD (Figure 3B). Based on the splicing coordinate, we found that AD event would occur in intron 1 and the density map of the GCA genome confirmed that there were fewer reads in sepsis (3,868 reads) compared to control (4,202 reads) in that coordinate, suggesting fewer AD events occurring in sepsis (Figure 3C). ELISA results showed that median plasma GCA concentration was statistically significantly higher in sepsis group compared to control (0 ng/mL vs. 0.81 ng/mL, p=0.04), in concordance with NMD pipeline prediction (Figure 3D). Weak positive correlation between RNA-Seq read counts and protein concentrations in sepsis (R=0.19, p=0.29) and moderate negative correlation in control (R=-0.41, p=0.42) were not statistically significant (Figure 3E).

**Fig. 3.**
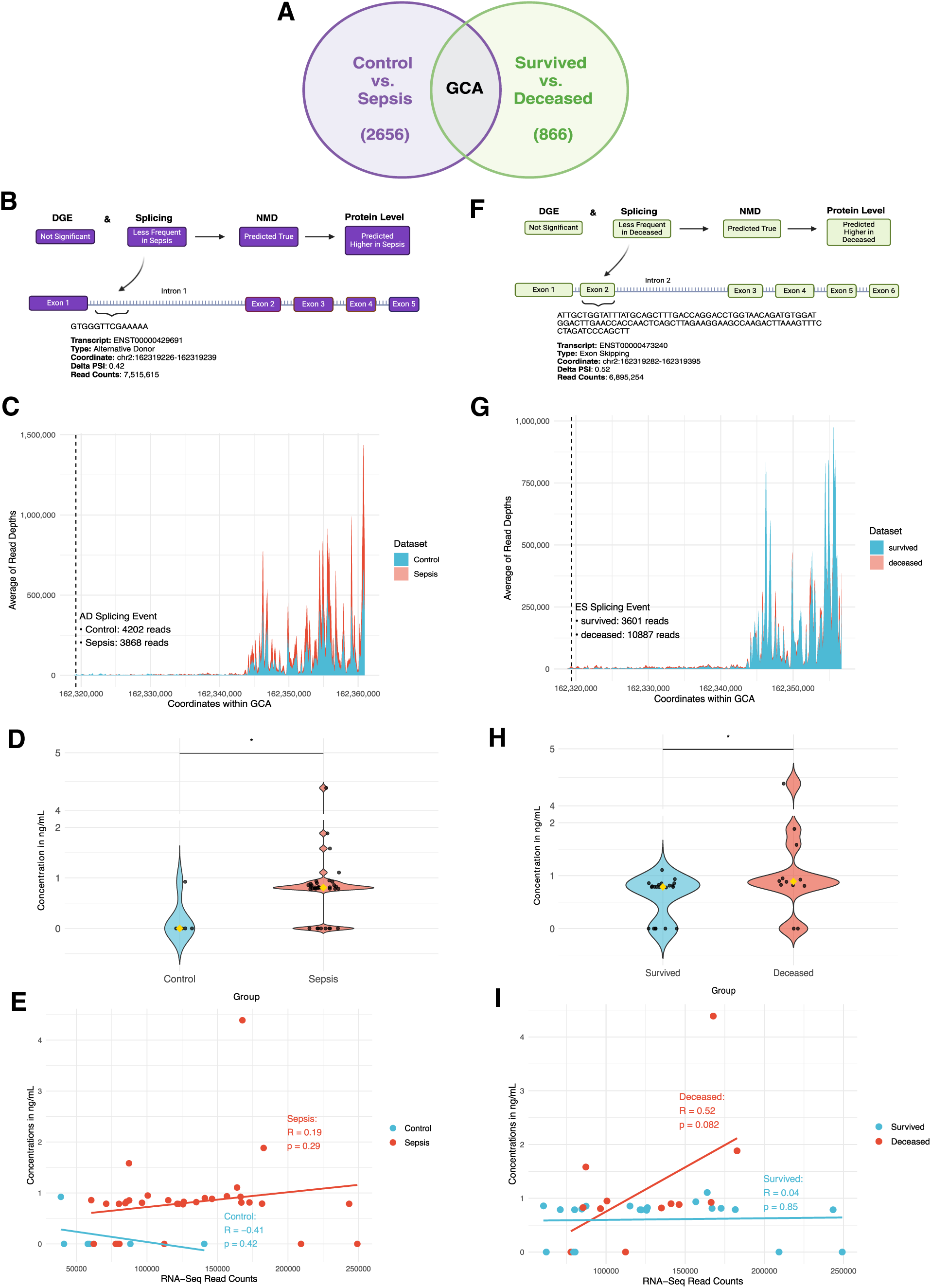
NMD pipeline prediction and proteomics data on plasma grancalcin (GCA) in control vs sepsis (Fig. 3B-E) and survived vs deceased groups (Fig. 3F-I). **(A)** Diagram describing GCA as the only gene with significant differential splicing in both control vs sepsis (total 2,656 significant differential splicing events) and survived vs deceased (total 866 significant differential splicing events) and with one of the highest absolute RNA-Seq read counts (ARC). Created in https://BioRender.com. **(B)** Diagram showing prediction on plasma GCA protein level based on DGE, splicing, and NMD data, along with the details on its significant differential splicing event in control vs sepsis. Created in https://BioRender.com. **(C)** Density map showing the average number of RNA-Seq reads per each 150 bp coordinate range of GCA genome. Dashed line represents the location of the alternative donor (AD) event (from Fig. 3B) and respective number of reads in control vs sepsis at the coordinates of AD event. **(D)** Violin plot showing the distribution and median ELISA protein concentrations of GCA in each sample in control vs sepsis. **(E)** Graph showing the correlation data between ELISA concentrations in ng/mL and RNA-Seq read counts of GCA in control vs sepsis. **(F)** Diagram showing prediction on plasma GCA protein level based on DGE, splicing, and NMD data, along with the details on its significant differential splicing event in survived vs deceased. Created in https://BioRender.com**. (G)** Density map showing the average number of RNA-Seq reads per each 150 bp coordinate range of GCA genome. Dashed line represents the location of the exon skipping (ES) event (from Fig. 3F) and respective number of reads in survived vs deceased at the coordinates of ES event. **(H)** Violin plot showing the distribution and median ELISA protein concentrations of GCA in each sample in survived vs deceased. **(I)** Graph showing the correlation data between ELISA concentrations in ng/mL and RNA-Seq read counts of GCA in survived vs deceased.

### Plasma GCA concentration is higher in deceased as predicted by NMD pipeline

Between survived and deceased group, GCA was also not significantly differentially expressed based on DGE, less frequently spliced in sepsis based on Whippet, and NMD predicted true based on the pipeline. Thus, higher protein level was predicted in deceased (Figure 3F). Based on the splicing coordinate, we found that ES event would occur in exon 2 and the density map of the GCA genome confirmed that there were higher reads in deceased (10,887 reads) compared to survived (3,601 reads) in that coordinate, supporting that there were fewer ES events in deceased group, thus more reads given more inclusion of exons (Figure 3G). ELISA results showed that median plasma GCA concentration was statistically significantly higher in deceased group compared to survived (0.79 ng/mL vs. 0.89 ng/mL, p=0.006), in concordance with NMD pipeline prediction (Figure 3H). While not statistically significant, higher RNA-Seq read counts demonstrated a trend toward moderate positive correlation with protein concentrations in deceased group (R=0.52, p=0.08) whereas it was not correlated in control (R=0.04, p=0.85) (Figure 3I).

## Discussion

We have developed a pipeline to predict how splicing events introduce PTCs into mRNA transcripts and induce PTC-dependent NMD. We showed that while more genes were upregulated in sepsis and downregulated in deceased based on DGE, a significantly higher proportion of transcripts were predicted to undergo NMD from splicing events in sepsis and deceased, which can influence downstream protein levels. Additionally, we have shown that NMD-F group can identify protein targets with potential novel roles in sepsis and that proteomics results on plasma GCA was consistent with the NMD pipeline prediction.

Our RNA-Seq data and downstream analyses have high biological and clinical relevance since they originated from the whole blood of 49 critically ill patients in the ICU with and without sepsis. This enabled an unbiased study of each patient’s transcriptome, augmented by the depth of 100 million reads that allowed a comprehensive DGE analysis and non-poly(A) tail-selection that incorporated splicing intermediates to facilitate AS studies. While DGE data showed upregulation of genes in sepsis and downregulation of genes in deceased patients, AS data showed transcripts were differentially spliced in nearly equivalent levels in both groups. This suggests AS may modulate downstream effects of over- or under-translation of proteins to maintain cellular homeostasis in diseased states (24). Unlike ES – most common form of splicing event (25) –, non-ES events(RI, AA, AD) were more common in sepsis and mortality, which indicates that non-ES events may exert a greater effect on sepsis and mortality status. To mitigate the loss of variance from the relative nature of RNA-Seq, we utilized ARC, quality control, and high sequencing depth (Supplementary Text). A significant difference between up- and down-regulated DGE results may be attributed to a higher log2FC threshold we used.

More than 90% rate of NMD seen across all groups can support that NMD is a conserved, ubiquitous subcellular process. A higher rate of NMD in sepsis and mortality is likely explained by acidosis, hypoxia, and fever or hypothermia that predispose cells to aberrant splicing (10–12) and splicing errors that necessitate higher rate of NMD (2). Moreover, AS-NMD mechanism may also be downregulating certain transcripts related to cellular stress signals (4) or affected by hypoxia (13) to regulate resources devoted to combating infection and preventing organ failure. Given the known coupling of AS-NMD in downregulating certain transcripts (5), a list of genes identified as NMD-F (Fig. 2H, 2I) demonstrate potential importance in sepsis and mortality, as evidenced by their involvement in nucleic acid and cell metabolism, signal transduction, inflammation, and response to stressor. Of note, all but 1 gene (PLAAT2) included in NMD-F group had statistically non-significant DGE results, thus were analyzed in GO terms as aggregates. Thus, NMD pipeline not only highlights AS-NMD interaction, but also identifies potentially significant protein targets in sepsis.

It is important to consider the intermediate transcriptional regulatory steps in gene expression that affect protein levels such as AS-NMD. To this end, we have shown that while DGE of GCA in sepsis and deceased was not significantly different, the NMD pipeline predicted higher GCA level in both conditions, which was corroborated by plasma protein ELISA. Given that RNA-Seq read counts and protein concentration data were not significantly correlated (Fig. 3E, 3I), other decay pathways (26, 27) and post-translational modifications may have affected the protein level, which may explain why plasma granulin was not consistent with the NMD pipeline prediction (Fig. S5-6). In addition, low detection rate in control group may be due to a lower proportion of GCA present in plasma samples from less severe inflammatory condition compared to sepsis. Regardless, NMD pipeline can potentially help discover protein targets previously unknown due to DGE studies alone. Since AS-NMD mechanism is highly conserved and essential throughout high eukaryotes (3), other fields can also utilize the NMD pipeline.

Our study does not establish definitive causality between AS-NMD machinery and gene expression in sepsis. However, we do provide a potential mechanistic insight of how altered splicing events can lead to NMD in sepsis via a novel computational pipeline. Future studies can include external validation of our computational work with a separate cohort of critically ill patients. From our sample size of around 50 patients, the depth of our RNA-Seq data covering at least 100 million RNA reads from each patient provided sufficient data points for our current investigation of predicting the rate of NMD from splicing events.

Overall, this study demonstrates that NMD pipeline can predict NMD from differential splicing events. In sepsis and deceased, there are higher rates of NMD from aberrant splicing requiring more frequent NMD or purposeful downregulation of certain genes. We also propose our NMD pipeline can be a component of gene expression studies, both in sepsis and other fields, to characterize post-transcriptomic gene products at levels detected under homeostasis and pathological conditions. Further, by investigating the role of AS-NMD in altered physiological states, we can better understand its potential pathological role and identify its impact on novel protein targets with potential diagnostic or therapeutic importance.

## Methods

### Study Approval

All patients or their appropriate surrogate provided informed consent as approved by the hospital Institutional Review Board (Approval #: 411616).

### Study Design

A single center, prospective study of critically ill patients with and without sepsis at an academic tertiary center was performed from 2021 to 2022.

Inclusion criteria for a diagnosis of sepsis were as follows: corresponding ICD-10 coding on admission, confirmed diagnosis of sepsis by attending physician, documentation of septic shock with evidence of hypo-perfusion(mean arterial pressure < 65, systolic blood pressure < 90 mmHg, lactate > 2 mmol/L after 30 mL/kg crystalloid bolus within 3 hours of identification), and the evidence of infection with at least one end-organ failure per SOFA score. Exclusion criteria included age less than 18 years old, pregnancy, incarceration, trauma within 30 days, immunocompromised conditions such as malignancy and pulmonary fibrosis, and known recent antibiotic use within 1 week before admission (11). Patients were enrolled from the medical intensive care unit upon admission or within 24 hours of developing sepsis in the ICU.

Whole blood samples were drawn in PAXgene tubes (Qiagen, Germantown, MD) on hospital day 0 then sent to Genewiz (South Plainfield, NJ) for RNA extraction, globin and ribosomal RNA depletion and deep RNA sequencing on Illumina HiSeq machines. Polyadenylic acid (poly A) tail selection was not performed to include all splicing intermediates and events. The sequencing provided 150 bp, paired end reads and at least 100 million reads per sample (28).

The raw sequencing data was assessed for quality control with FastQC (29) then aligned to the most recent assembly (GCF_000001405.40) of Genome Reference Consortium Human Build 38 (GRCh38) (30) with STAR aligner (31). Reads that aligned to GRCh38 (“mapped” reads) were separated from “unmapped” reads. The mapped reads from control vs sepsis and survived vs deceased group then underwent analysis for DGE and AS.

For DGE, featureCounts (32) was used to yield raw absolute read counts (ARC), then DESeq2 package from Bioconductor (33) helped identify differentially expressed genes. Statistical significance was determined as adjusted p<0.05 and |log2foldchange|>2 (12). For AS, Whippet (23) was used to compare exon skipping (ES), retained intron (RI), alternative donor (AD) and alternative acceptor (AA) events. Statistical significance was determined by probability>=0.9 and |DeltaPsi|>0.1 per Whippet documentation (34). For NMD pipeline, we leveraged Whippet AS output data to develop a computational approach to examine the rate of NMD, number of PTCs generated, stratification by splicing subtype, and GO Enrichment Analysis on splicing events with predicted false NMD (Supplementary Text).

We performed enzyme-linked immunosorbent assay (ELISA) of grancalcin (GCA) (MyBioSource, Catalog #: <u>MBS2709681</u>) and granulin (GRN) (Invitrogen, Catalog #: EH367RB) with adherence to commercially available ELISA protocols from respective manuals. Two replicates of each sample were used to calculate an average of the two optical density (OD) values to compute final concentrations. Concentrations of any OD below the detectable range indicated by the manufacturer were imputed as zero.

All computational and statistical analyses were done in R (35) and command line *bash, awk, grep* script. For continuous variables, either t-test or Wilcoxon test was used based on Shapiro-Wilk normality test. For categorical variables, chi-square test with or without Yates’ correction for continuity was used based on the sample sizes.

## Data Availability

All data produced in the present study are available upon reasonable request to the authors.

## Data availability

Our data sets and code utilized and developed for the computational pipeline will be available in the appropriate public repository.

## Authors’ Contributions

Conceptualization: JS, AMF, SFM; Methodology: JS, AMF, WGF, ER, MML, SFM; Investigation: JS, AMF, SFM; Visualization: JS, AMF, BEA, KKL, ER, SFM; Funding acquisition: AA, MC, WGF, MML, GJN, SFM; Project administration: JS, AMF, SFM; Supervision: AFM, BEA, AA, WGF, GJN, SFM; Writing – original draft: JS; Writing – review & editing: JS, AMF, BEA, AA, WGF, KKL, ER, GJN, SFM

## Acknowledgments

We thank R. Zhao of the Division of Surgical Research in the Department of Surgery at Rhode Island Hospital for valued instructions with proteomics work; Biorender for allowing the creation of several subfigure flowcharts.

## Supplementary Materials

Figs. S1 to S6

Tables S1 to S12

Supplementary Text

