## Supplementary Materials for "Predicting Nonsense-mediated mRNA Decay from Splicing Events in Sepsis using RNA-Sequencing Data"

**This file includes:**

Supplementary Figures S1-6
Supplementary Tables S1-12
Supplementary Text

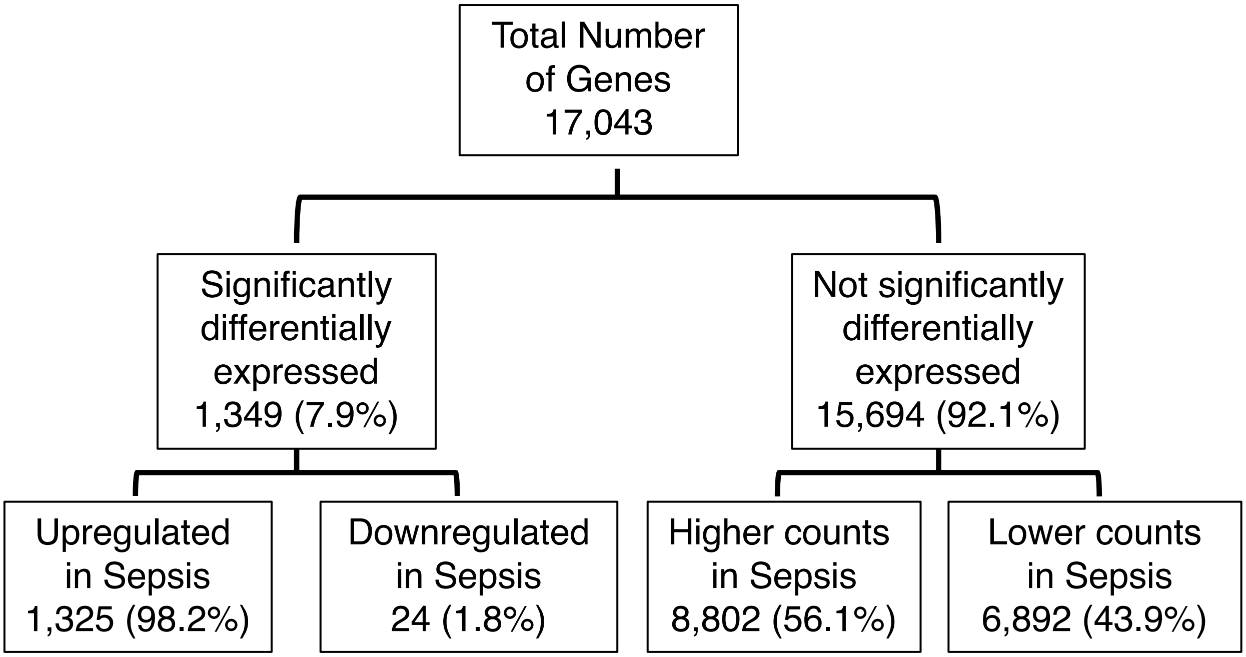


**Fig. S1.**
Differential gene expression (DGE) datapoints for the volcano plot in control vs sepsis (Fig. 1B).

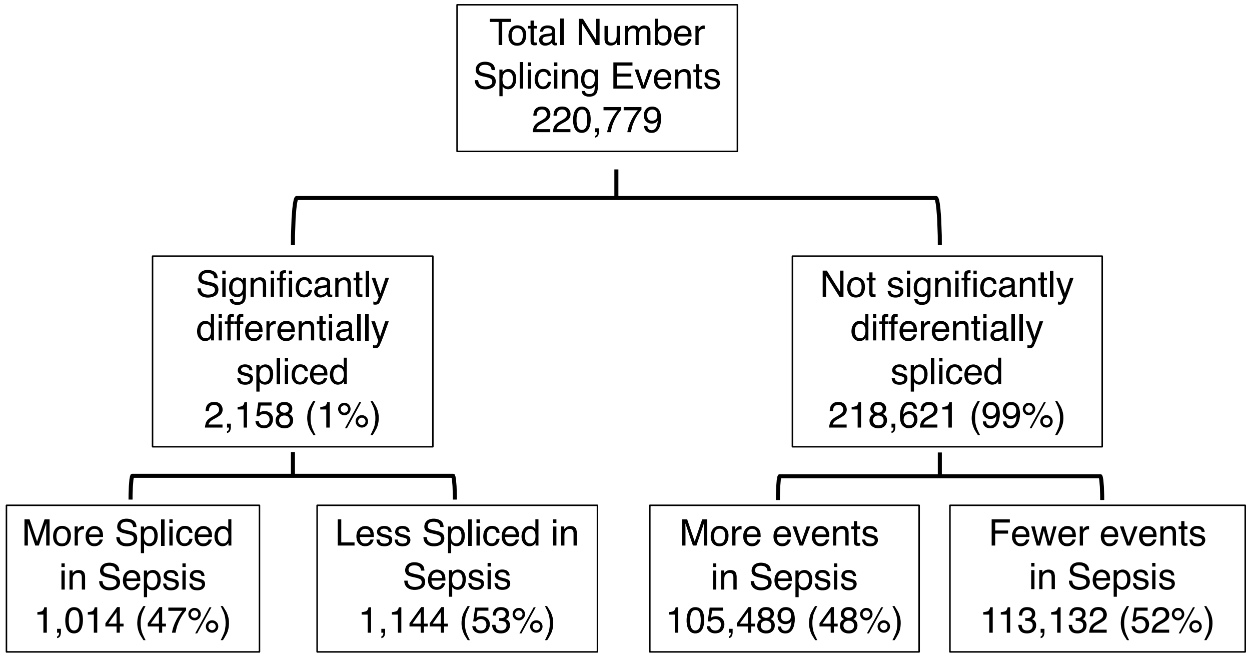


**Fig. S2.**
Differential splicing analysis datapoints for the volcano plot in control vs sepsis (Fig. 1C).

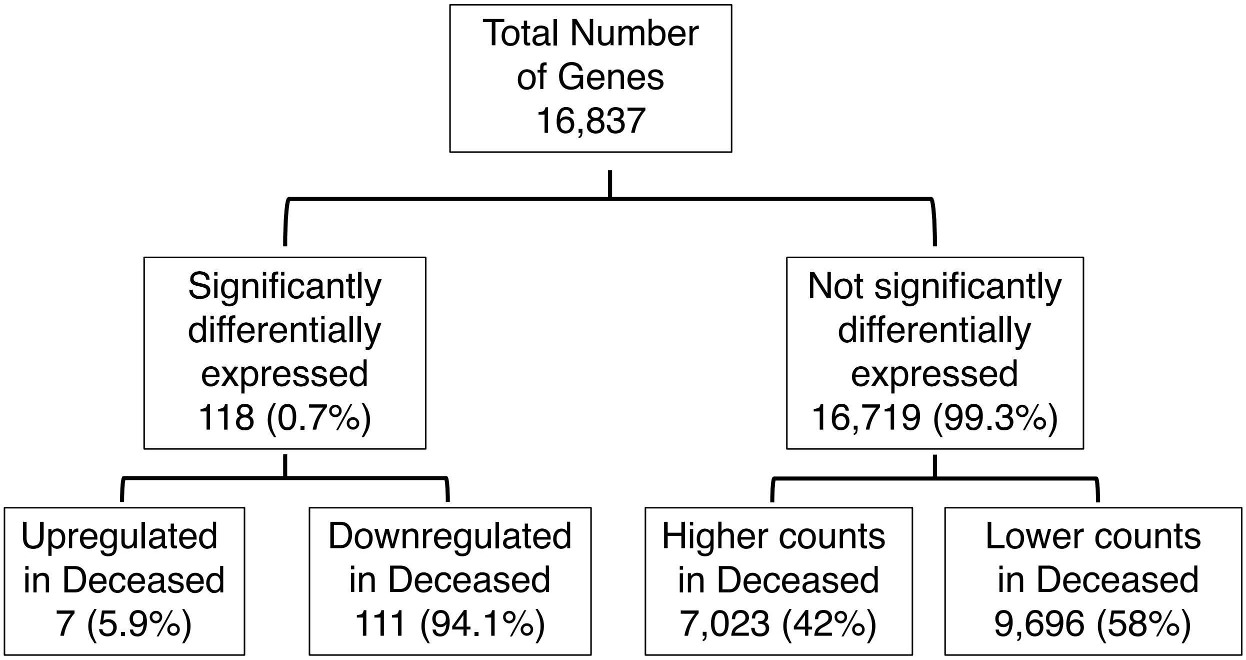


**Fig. S3.**

30 Differential gene expression (DGE) datapoints for the volcano plot in survived vs deceased (Fig. 1F).

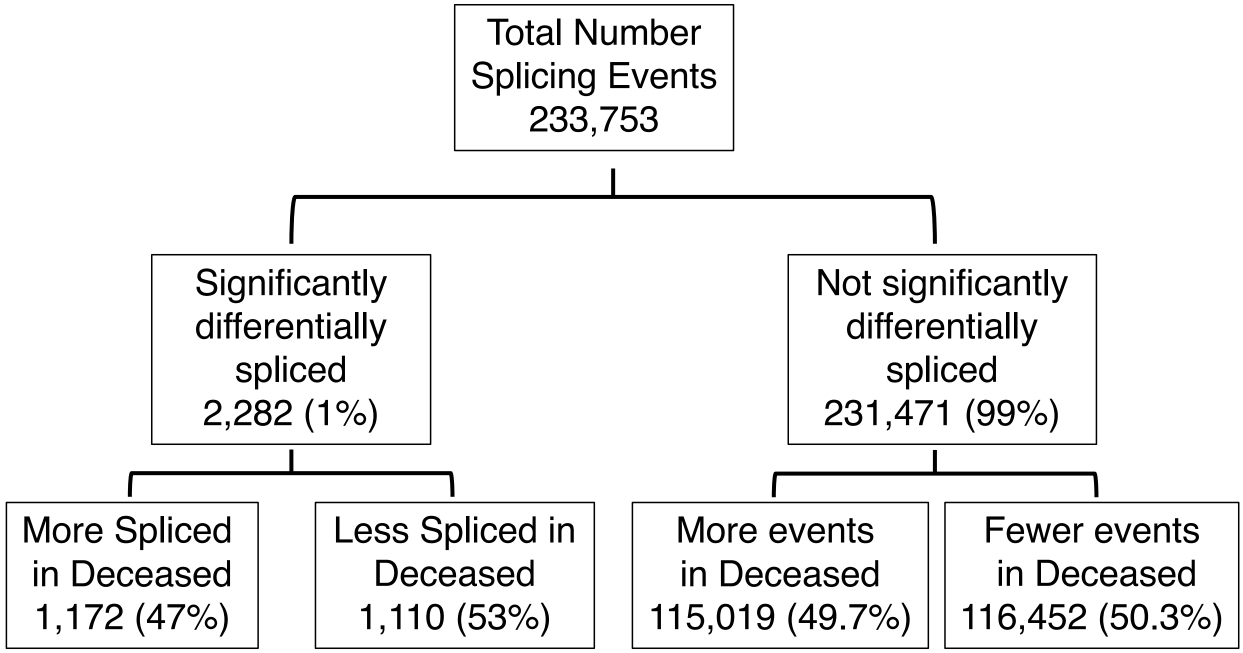


**Fig. S4.**
Differential splicing analysis datapoints for the volcano plot in survived vs deceased (Fig. 1G).

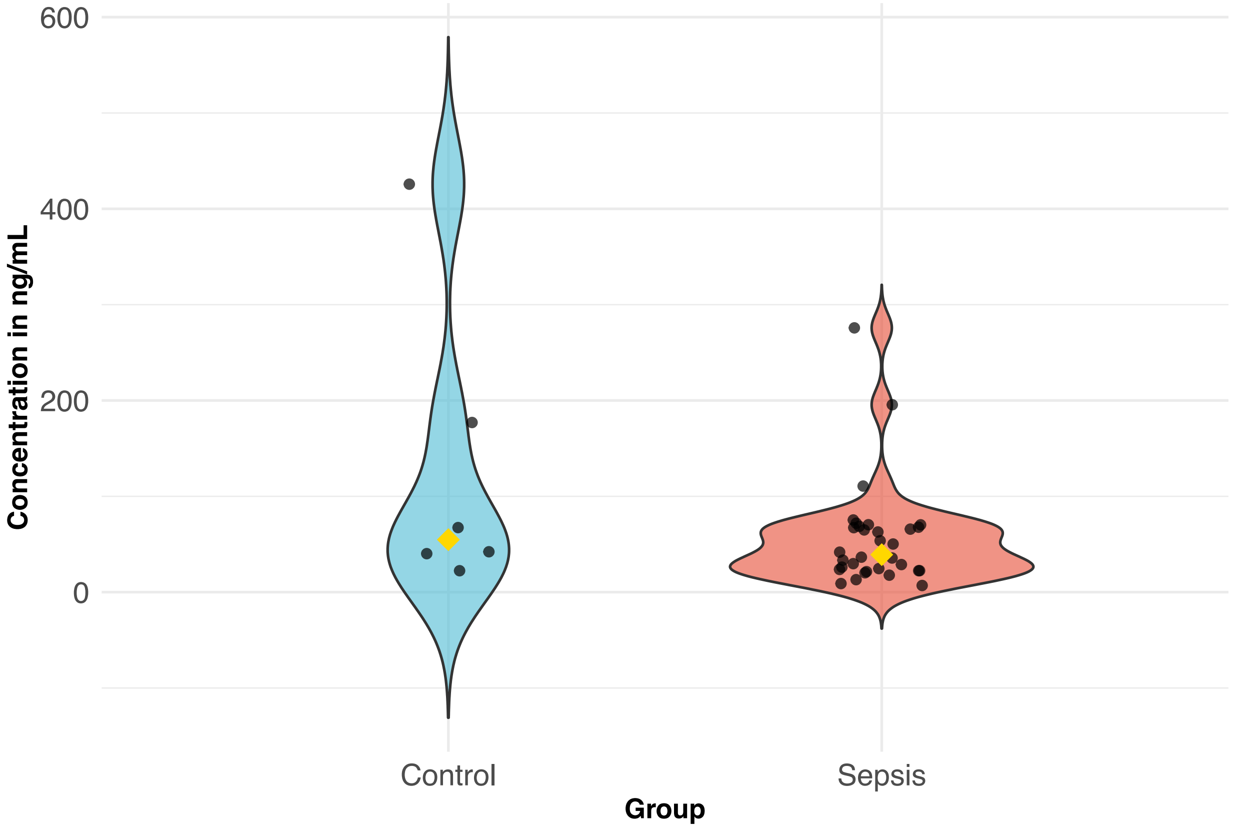


**Fig. S5.**

Violin plot showing the distribution and median ELISA protein concentrations of plasma granulin in each sample in control vs sepsis.

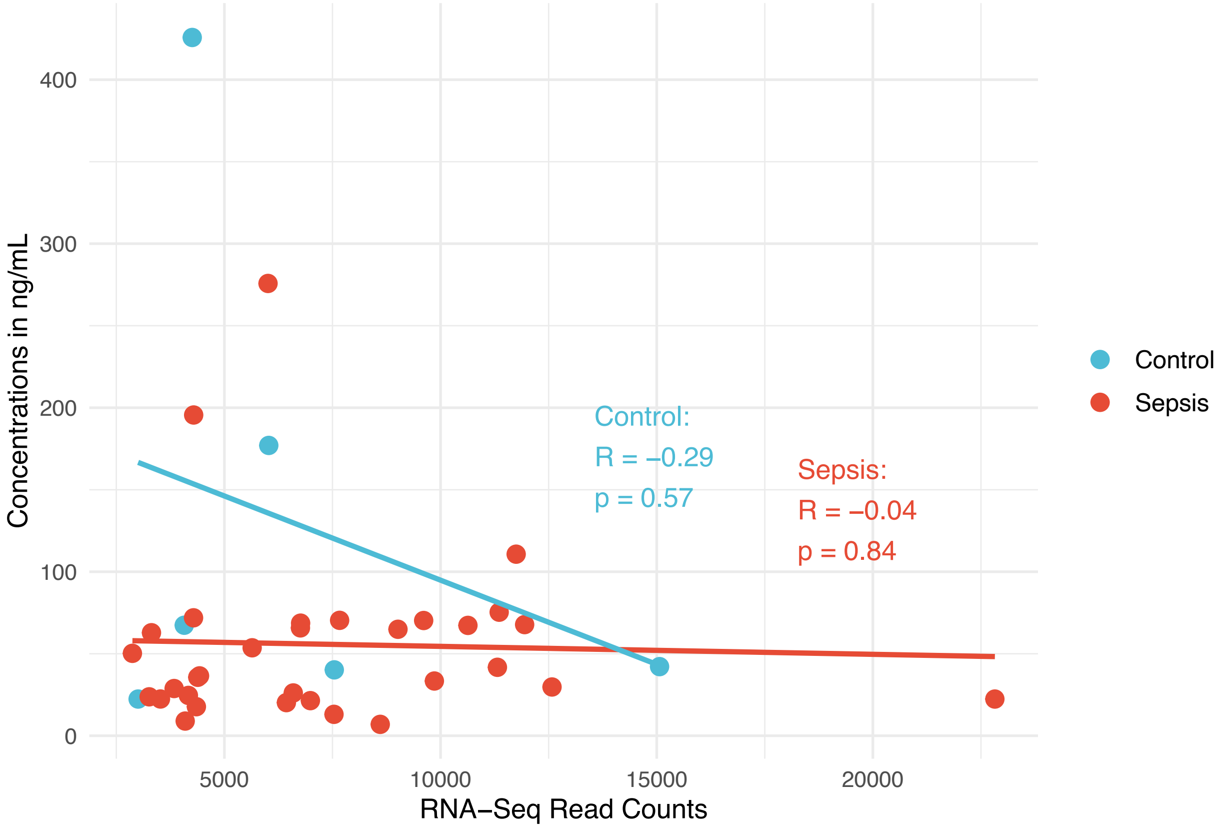


**Fig. S6.**

Graph showing the correlation data between ELISA concentrations in ng/mL and RNA-Seq read counts of plasma granulin in control vs sepsis.

**Table S1.**

Percentage breakdown of each splicing event categorized as “Splicing” and “Transcription” groups in control vs sepsis (Fig. 1D).

| Control | | Sepsis | |
|---|---|---|---|
| **Splicing Events** | **Percentages (51.9%)** | **Splicing Events** | **Percentages (45%)** |
| Exon Skipping | 76.3% | Exon Skipping | 44.7% |
| Retained Intron | 9.5% | Retained Intron | 19.5% |
| Alternative Donor | 8.1% | Alternative Donor | 18% |
| Alternative Acceptor | 6.1% | Alternative Acceptor | 17.8% |
| **Transcription** | **Percentages (48.1%)** | **Transcription** | **Percentages (55%)** |
| Transcription Start | 31.7% | Transcription Start | 46.7% |
| Transcription End | 62.8% | Transcription End | 48.9% |
| Alternative First | 3.9% | Alternative First | 3% |
| Alternative Last | 1.6% | Alternative Last | 1.4% |

**Table S2.**

Frequency (in percentage) of each splicing event subtype in control vs sepsis (Fig. 1E).

| **Splicing Events** | **Control** | **Sepsis** | **p value** |
|---|---|---|---|
| Exon Skipping | 76.3% | 44.7% | < 0.001 |
| Retained Intron | 9.5% | 19.5% | < 0.001 |
| Alternative Acceptor | 8.1% | 18% | < 0.001 |
| Alternative Donor | 6.1% | 17.8% | < 0.001 |

**Table S3.**

Percentage breakdown of each splicing event categorized as "Splicing" and "Transcription" groups in control vs sepsis (Fig. 1H).

| Control | | Sepsis | |
|---|---|---|---|
| **Splicing Events** | **Percentages (50.5%)** | **Splicing Events** | **Percentages (37.9%)** |
| Exon Skipping | 76.1% | Exon Skipping | 48.5% |
| Retained Intron | 9.5% | Retained Intron | 21.7% |
| Alternative Donor | 8.2% | Alternative Donor | 15.6% |
| Alternative Acceptor | 6.2% | Alternative Acceptor | 14.2% |
| **Transcription** | **Percentages (49.5%)** | **Transcription** | **Percentages (62.1%)** |
| Transcription Start | 33.6% | Transcription Start | 51% |
| Transcription End | 61% | Transcription End | 46% |
| Alternative First | 1.6% | Alternative First | 1.8% |
| Alternative Last | 3.8% | Alternative Last | 1.2% |

**Table S4.**

Frequency (in percentage) of each splicing event subtype in survived vs deceased (Fig. 1I).

| Splicing Events | Survived | Deceased | p value |
| --- | --- | --- | --- |
| Exon Skipping | 76.1% | 48.5% | < 0.001 |
| Retained Intron | 9.5% | 21.7% | < 0.001 |
| Alternative Acceptor | 8.2% | 15.6% | < 0.001 |
| Alternative Donor | 6.2% | 14.2% | < 0.001 |

**Table S5.**

Percentage of splicing events predicted to induce NMD in control vs sepsis (top) and the percentage of predicted NMD stratified by splicing subtypes (bottom) (Fig. 2B). Total Canonical refers to ENSEMBL canonical transcripts that NMD pipeline is built to process.

| **Percentage of NMD in Control vs Sepsis groups** | | | |
|---|---|---|---|
| | Control | Sepsis | p value |
| Total Splicing Events | 114,487 | 994 | |
| Total Canonical | 95,430 | 676 | -- |
| Predicted NMD True | 86,581 (90.7%) | 631 (93.3%) | 0.03 |
| Predicted NMD False | 8,849 (9.3%) | 45 (6.7%) | -- |
| **Percentage of NMD in Control vs Sepsis groups per subtype** | | | |
| | NMD True in Control | NMD True in Sepsis | p value |
| Exon Skipping | 69,732 (89.7%) | 245 (92.4%) | 0.16 |
| Retained Intron | 9,272 (92.4%) | 168 (97%) | 0.61 |
| Alternative Acceptor | 3,750 (92.8%) | 102 (95.3%) | 0.43 |
| Alternative Donor | 3,827 (94.4%) | 107 (89.2%) | 0.03 |

**Table S6.**

Proportion of splicing events of transcripts predicted to cause NMD per each splicing subtype in control vs sepsis (Fig. 2C).

| Splicing Events | Control | Sepsis | p value |
|---|---|---|---|
| Exon Skipping | 80.5% | 39.4 % | < 0.001 |
| Retained Intron | 10.7% | 27% | < 0.001 |
| Alternative Acceptor | 4.3% | 16.4% | < 0.001 |
| Alternative Donor | 4.4% | 17.2% | < 0.001 |

**Table S7.**

Total and median number of premature termination codons (PTCs) generated per splicing subtype in control vs sepsis (Fig. 2D).

| | **Control** | | | **Sepsis** | | | |
|---|---|---|---|---|---|---|---|
| | Total PTCs generated per subtype | Total Events per subtype | Median PTCs per Subtype | Total PTCs generated per subtype | Total Events per Subtype | Median PTCs per Subtype | p value |
| ES | 3,873,406 | 69,732 | 37 | 17,950 | 245 | 36 | 0.51 |
| RI | 355,273 | 9,272 | 23 | 6,416 | 168 | 21 | 0.68 |
| AA | 150,304 | 3,750 | 24 | 3,738 | 102 | 24 | 0.74 |
| AD | 174,121 | 3,827 | 30 | 5,029 | 107 | 35 | 0.43 |

**Table S8.**

Percentage of splicing events predicted to induce NMD in survived vs deceased (top) and the percentage of predicted NMD stratified by splicing subtypes (bottom) (Fig. 2E). Total Canonical refers to ENSEMBL canonical transcripts that NMD pipeline is built to process.

| **Percentage of NMD in Survived versus Deceased groups** | | | |
|---|---|---|---|
| | Survived | Deceased | p value |
| Total Splicing Events | 118,152 | 866 | -- |
| Total Canonical | 98,177 | 579 | -- |
| Predicted NMD True | 89,095 (90.8%) | 540 (93.3%) | 0.04 |
| Predicted NMD False | 9,082 (9.3%) | 39 (6.7%) | -- |
| **Percentage of NMD in Survived versus Deceased groups per subtype** | | | |
| | Survived | Deceased | p value |
| Exon Skipping | 71,698 (89.7%) | 216 (89.6%) | 1 |
| Retained Intron | 9,495 (96.9%) | 167 (96%) | 0.64 |
| Alternative Acceptor | 3,856 (92.7%) | 75 (94.9%) | 0.59 |
| Alternative Donor | 4,046 (94.4%) | 82 (96.5%) | 0.56 |

**Table S9.**

Proportion of splicing events of transcripts predicted to cause NMD per each splicing subtype in survived vs deceased (Fig. 2F).

| **Splicing Events** | **Survived** | **Deceased** | **p value** |
|---|---|---|---|
| Exon Skipping | 80.5% | 40 % | < 0.001 |
| Retained Intron | 10.7% | 30.9% | < 0.001 |
| Alternative Acceptor | 4.3% | 13.9% | < 0.001 |
| Alternative Donor | 4.5% | 15.2% | < 0.001 |

**Table S10.**

Total and median number of premature termination codons (PTCs) generated per splicing subtype in survived vs deceased (Fig. 2G).

| | **Survived** | | | **Deceased** | | | |
|---|---|---|---|---|---|---|---|
| | Total PTCs generated per subtype | Total Events per subtype | Median PTCs per Subtype | Total PTCs generated per subtype | Total Events per Subtype | Median PTCs per Subtype | p value |
| ES | 3,985,343 | 71,698 | 37 | 10,793 | 216 | 39.5 | 0.73 |
| RI | 362,735 | 9,495 | 23 | 5,070 | 167 | 19 | 0.07 |
| AA | 154,298 | 3,856 | 24 | 2,580 | 75 | 18 | 0.47 |
| AD | 186,108 | 4,046 | 31 | 3,472 | 82 | 28.5 | 0.58 |

**Table S11.**

GO Enrichment Analysis results for all transcripts with splicing events not expected to undergo NMD with $p < 0.01$ in control vs sepsis (Fig. 2H).

| ID | Description | Gene Ratio | p value | p adjust | Gene ID | Count |
|---|---|---|---|---|---|---|
| GO:0010800 | positive regulation of peptidyl-threonine phosphorylation | 3/40 | 0.00003 | 0.03051 | EGF/PLK1/STOX1 | 3 |
| GO:0010799 | regulation of peptidyl-threonine phosphorylation | 3/40 | 0.00011 | 0.06024 | EGF/PLK1/STOX1 | 3 |
| GO:0010288 | response to lead ion | 2/40 | 0.00090 | 0.24087 | PPP5C/BACE1 | 2 |
| GO:0009162 | deoxyribonucleoside monophosphate metabolic process | 2/40 | 0.00108 | 0.24087 | TK1/NT5C | 2 |
| GO:0018107 | peptidyl-threonine phosphorylation | 3/40 | 0.00110 | 0.24087 | EGF/PLK1/STOX1 | 3 |
| GO:0018210 | peptidyl-threonine modification | 3/40 | 0.00146 | 0.25742 | EGF/PLK1/STOX1 | 3 |
| GO:1905332 | positive regulation of morphogenesis of an epithelium | 2/40 | 0.00263 | 0.25742 | EGF/STOX1 | 2 |
| GO:0007143 | female meiotic nuclear division | 2/40 | 0.00308 | 0.25742 | PLK1/HSF2BP | 2 |
| GO:0043124 | negative regulation of canonical NF-kappaB signal transduction | 2/40 | 0.00811 | 0.25742 | TRIM59/TSPAN6 | 2 |
| GO:1905330 | regulation of morphogenesis of an epithelium | 2/40 | 0.00836 | 0.25742 | EGF/STOX1 | 2 |
| GO:0006399 | tRNA metabolic process | 3/40 | 0.00890 | 0.25742 | GATB/EXOSC7/PUSL1 | 3 |

**Table S12.**

GO Enrichment Analysis results for all transcripts with splicing events not expected to undergo NMD with $p < 0.01$ in survived vs deceased (Fig. 2I).

| ID | Description | Gene Ratio | p value | p adjust | Gene ID | Count |
|---|---|---|---|---|---|---|
| GO:0045471 | response to ethanol | 3/31 | 0.00104 | 0.21654 | SDF4/NQO1/RPL15 | 3 |
| GO:0009411 | response to UV | 3/31 | 0.00202 | 0.21654 | MAP4K3/SDF4/CCND1 | 3 |
| GO:0019674 | NAD metabolic process | 2/31 | 0.00493 | 0.21654 | NQO1/QPRT | 2 |
| GO:0044772 | mitotic cell cycle phase transition | 4/31 | 0.00606 | 0.21654 | ACTB/KDM8/CCND1/BIRC5 | 4 |
| GO:0046470 | phosphatidylcholine metabolic process | 2/31 | 0.00689 | 0.21654 | PEMT/PLAAT2 | 2 |
| GO:0000079 | regulation of cyclin-dependent protein serine/threonine kinase activity | 2/31 | 0.00724 | 0.21654 | ACTB/CCND1 | 2 |
| GO:1904029 | regulation of cyclin-dependent protein kinase activity | 2/31 | 0.00779 | 0.21654 | ACTB/CCND1 | 2 |
| GO:0097305 | response to alcohol | 3/31 | 0.00863 | 0.21654 | SDF4/NQO1/RPL15 | 3 |
| GO:0030071 | regulation of mitotic metaphase/anaphase transition | 2/31 | 0.00954 | 0.21654 | ACTB/BIRC5 | 2 |

### Supplementary Text

Supplementary Materials & Methods

For DGE, raw absolute read counts (ARC) were utilized to account for the relative nature of RNA-Seq so that the loss of variance from normalization can be mitigated, along with FastQC, DESeq2 built-in algorithm, and high sequencing depth of 100 million reads.

The splicing events identified by Whippet were then categorized as "Splicing" and "Transcription-related" groups to differentiate splicing events from transcription-related events in preparation for NMD processing. Of note, Whippet output includes core exon (CE) events which refer to exons involved in exon skipping (30), thus we used the nomenclature ES for clearer delineation.

When performing comparisons of splicing events in control vs sepsis and survived vs deceased, we utilized all differential spliced events vs significant differential splicing events. The reason for this was that Whippet outputs differential splicing event data of sepsis or deceased group with respect to control or survived group. Thus, all splicing events would effectively represent the control and survived group as a baseline for comparison and significant splicing events would effectively represent sepsis and deceased group.

We wrote a code in R Script that utilizes splicing event information (e.g. Ensembl gene id, splicing subtype, splicing event coordinate, positive or negative strand of the transcript with the splicing event) to predict whether each splicing event would generate premature termination codons (PTCs) thereby predicted to induce NMD. Thus, the outputs of our code include predicted frame of codons of each transcript (based on UCSC genome browser), the number of PTCs to be introduced by each splicing event for each possible frame, and a true vs false result of whether NMD would be induced based on whether PTCs would be generated in a predicted frame. Of note, the scope of our code was to predict PTC generation that elicits PTC-dependent NMD for PTCs 50-55 bp upstream of final exon junction.

Of note, our NMD pipeline was designed to process all splicing events designated as ENSEMBL canonical transcripts. The rationale was two-fold: first, Whippet output does not designate Ensembl Transcript ID thus there needed to be an evidence-based method to select 1 representative transcript corresponding to each Ensembl gene id; second, ENSEMBL canonical transcripts are the most representative transcripts that balance the highest coverage of conserved exons, expression, and consistency with other resources such as NCBI thus was chosen as an input to our NMD pipeline. We show further data on the canonical transcripts in Table S5 and S10.

We selected grancalcin (GCA) as our main protein target for 3 reasons: first, it was one of the most abundant targets found in our analysis based on RNA-Seq read counts, which indicate its potential significance in sepsis; second, it was the only target with significant differential splicing events in both control vs sepsis and survived vs deceased, which suggest it may be important in both diagnosis and prognosis of sepsis. Plasma patient samples were used to test the targets given their availability in our lab and their practicality as a clinical test. For this reason, we also selected plasma granulin (GRN) as an additional protein target to test for control vs sepsis.

Coding details for NMD pipeline

Once Whippet output data were imported to the R environment, relevant inputs (e.g. Ensembl gene id, splicing event type, splicing event coordinate, and strand) were utilized to access the corresponding nucleotide sequences of the splicing event from UCSC Genome Browser. Then exon coordinates were extracted and intron coordinates were deduced. Then mature mRNA transcript sequences were modeled based on each type of splicing event. For instance, if exon skipping event of exon 2 occurred, then the mature transcript sequence was coded to output exon

1, exon 3, exon 4, so forth. Code was written to account for each type of splicing event including exon skipping (ES), retained intron (RI), alternative donor (AD), and alternative acceptor (AA). Then, the predicted frame was calculated based on the codon information listed on UCSC Genome Browser and another code was written to search for 3 conventional stop codons (TAA, TAG, TGA) in the mature transcript with alternative splicing event. Finally, NMD was predicted to occur if at least 1 stop codon was generated in the predicted frame 50-55 base pairs upstream of final exon junction, which would qualify as premature termination codons (PTC). Thus, for every splicing event of a canonical transcript extracted from Whippet data, our NMD pipeline NMD yields outputs consisting of predicted frame, number of PTCs for each frame, and NMD true or false.
